# Genetic analysis of asymptomatic antinuclear antibody production

**DOI:** 10.1101/2024.08.29.24312782

**Authors:** Mehmet Hocaoglu, Desiré Casares-Marfil, Amr H. Sawalha

## Abstract

**Objective:** Antinuclear antibodies (ANA) are detected in up to 14% of the population and the majority of individuals with ANA are asymptomatic. The literature on the genetic contribution to asymptomatic ANA positivity in the population is limited. In this study, we aimed to perform a multi-ancestry genome-wide association study (GWAS) of asymptomatic ANA positivity.

**Methods:** Asymptomatic ANA positive and negative individuals from the All of Us Research Program were included in this study, selecting those with an ANA test by immunofluorescence and no evidence of autoimmune disease. Imputation was performed and a multi-ancestry meta-analysis including approximately 6 million single-nucleotide polymorphisms (SNPs) was conducted. Genome-wide SNP based heritability was estimated using the GCTA software. A cumulative genetic risk score for lupus was constructed using previously reported genome-wide significant loci.

**Results:** 1,955 asymptomatic ANA positive and 3,634 asymptomatic ANA negative individuals were included across three genetic ancestries. The multi-ancestry meta-analysis revealed SNPs with a suggestive association (p-value < 1×10^−5^) across 8 different loci, but no genome-wide significant loci were identified. A gene variant upstream of *HLA-DQB1* (rs17211748, *P* = 1.4×10^-6^, OR = 0.82, 95% CI 0.76-0.89) showed the most significant association. The heritability of asymptomatic ANA positivity was estimated to be 24.9%. Asymptomatic ANA positive individuals did not exhibit increased cumulative genetic risk for lupus compared to ANA negative individuals.

**Conclusion:** ANA production is not associated with significant genetic risk and is primarily determined by non-genetic, likely environmental, factors.

## Introduction

Antinuclear antibodies (ANA) are associated with several immune-mediated diseases such as systemic lupus erythematosus (SLE), systemic sclerosis, Sjögren’s syndrome, and others (1). The population prevalence of ANA positivity in the United States was estimated to be 13.8% (95% confidence interval 12.2–15.5%), with evidence suggesting rising prevalence of autoimmunity over recent decades (2, 3). Environmental factors such as body-mass index and parity status have been shown to be associated with ANA positivity (2, 4), but the role of genetic factors in ANA production is less defined. Only one genome-wide association study (GWAS) has been performed, showing the association of a variant near the *HLA-DRA* gene with ANA positivity in individuals of Japanese ancestry (5).

Previous research showed that ANA production precedes the onset of clinical symptoms in SLE, suggesting that asymptomatic ANA positivity might be an intermediate state on the pathway to the development of clinical autoimmunity (6). However, most individuals with positive ANA do not develop any immune-mediated diseases (7). As autoimmune diseases are complex conditions arising from the interplay of genetic and environmental factors (8), understanding the genetic factors associated with asymptomatic ANA production could provide insights into the early pathways of autoimmunity development and differentiate them from the terminal pathways that lead to clinical pathology. In this study, we aimed to assess the role of genetic susceptibility in ANA production in a large multi-ancestral cohort.

## Methods

### Study population and design

The All of Us Research Program is a cohort study aiming to enroll 1,000,000 participants with genomic data linked with environmental surveys and electronic health records (EHR). As of the latest data release (version 7), All of Us included 312,925 and 245,388 individuals with genotyping and whole-genome sequencing data, respectively (9). For the GWAS analysis, we selected participants who had genotyping data available and a positive ANA test in their EHR by immunofluorescence (IF) as defined by the performing lab or if titer data are available, titer greater than 1:80 (**Supplementary List 1**). We excluded individuals who had one positive ANA test but subsequently had a negative test by IF or ELISA. We did not include individuals who only had a positive ANA by ELISA in this study (n = 225). Individuals with any clinical code for autoimmune diseases or prescription for an immunosuppressive medication in their EHR from both the ANA positive and ANA negative cohorts were also excluded (**Supplementary Figure 1, Supplementary List 2**). We performed a sensitivity analysis by restricting the analyses to individuals who had titer data available and had an ANA titer greater than 1:160.

### Quality controls, genetic ancestry assignment, and imputation

The individuals participating in the All of Us underwent genotyping with the Infinium Global Diversity Array-8 (Illumina). Variants with minor allele frequency (MAF) less than 1%, those deviated from Hardy-Weinberg equilibrium (HWE) in cases and controls (*P* < 1×10^−3^), and variants with high missingness rates (≥ 0.03) were removed. In addition, A/T-C/G SNPs with a MAF greater than 0.4 were removed prior to imputation to avoid errors with the imputation process. Individuals with poor genotyping rates (< 97%), cryptic relatedness as defined by their identity by descent proportion (PI-HAT > 0.4) or missing sex information were excluded. Sex chromosomes were not analyzed in this study. The genetic ancestry of the participants was assigned by intersecting their genetic information with the 1000 Genomes Project populations and calculating the first ten principal components (PCs) of the genotype-covariance matrix.

Subsequently, a random forest classifier was trained to assign to each participant included in our study to one of the following genetic ancestries per the standard superpopulations from the 1000 Genomes Project: European (EUR), Admixed American (AMR), East Asian (EAS), South Asian (SAS), and African (AFR) (10). As the sample size for EAS and SAS genetic ancestries were limited (n =115 and n = 52, respectively), they were excluded from further analyses.

Individuals who are 6 standard deviation away from their respective genetic ancestry centroid were considered outliers and were removed from further analyses. Principal component analysis was performed using the R package *smartsnp* which uses the EIGENSTRAT method (11, 12). Phasing and imputation were performed using Eagle2 and minimac4, respectively (13, 14). All populations from the 1000 Genomes Project phase 3 were used as reference panel for imputation and phasing. Imputed SNPs with an imputation quality metric less than RSQ 0.9 and a minor allele frequency less than 1% were removed from further analyses. Imputation procedures were performed for each genetic ancestry separately. Quality controls and statistical tests were performed using PLINK v1.9 (15).

### Statistical Analysis

GWAS was performed through logistic regression on each genetic ancestry separately using the first 5 PCs as covariates, followed by a meta-analysis using the inverse-variance method.

Variants with no evidence of heterogeneity were considered under fixed-effects model (Cochran’s Q test p-value > 0.1 and heterogeneity index < 50%) while the rest were considered under a random-effects model (Cochran’s Q test p-value ≤ 0.1 and heterogeneity index ≥ 50%). Only variants studied in more than one genetic ancestry GWAS were included in the meta-analysis. The genome-wide significance level was established at a p-value < 5×10^−8^ and the suggestive association threshold was set at a p-value < 1×10^−5^. In addition, the most significant association identified in the meta-analysis was assessed in SLE, compared to ANA positive and ANA negative cohorts, using logistic regression in individual ancestries followed by trans-ancestry meta-analysis as described for the main GWAS. For this analysis, we selected SLE cases in the All of Us database with genotyping data by including individuals who had at least two SLE clinical codes 30 days apart and at least two SLE related medication prescription in their EHR (**Supplementary List 3**).

### Genome-wide SNP-based heritability estimation

We estimated the genome-wide SNP heritability based on the liability scale for ANA positivity using the GCTA software (16), which utilizes a linkage disequilibrium (LD) and MAF stratified genome-based restricted maximum likelihood approach to correct for LD bias. For liability calculations, we assumed the prevalence of asymptomatic ANA positivity in the population to be 15% and used the first 5 PCs as covariates. This analysis was performed only in individuals of European ancestry (asymptomatic ANA positive n = 1,236, asymptomatic ANA negative n = 2,406) as the sample size required to provide reliable estimates (SD < 0.1) could only be achieved with total sample sizes greater than 3,000 individuals (17).

### Cumulative genetic risk score in SLE

To assess the variability of SLE genetic risk between SLE patients and asymptomatic ANA positive and negative individuals, a cumulative genetic risk score (GRS) was calculated. For this analysis, we included individuals with available whole-genome sequencing data in order to extract all genetic data needed to accurately calculate a GRS (**Supplementary Table 1**). We calculated an SLE GRS using 183 genetic susceptibility loci previously reported with a GWAS level of significance for SLE (18). Of the 183 SNPs we excluded five SNPs in sex chromosomes and one additional SNP (rs11845506) due to a case MAF < 1% in the study where it was reported, leaving 177 variants for GRS calculation (**Supplementary Table 2**). The GRS was estimated by multiplying the natural logarithm of the previously reported SLE association odds ratio (OR) for each SNP representing each locus by the risk allele counts of the corresponding SNPs as we previously described (19). We compared the mean GRS of each group with the other two groups using Tukey’s multiple comparison tests.

## Results

A total of 1,955 asymptomatic ANA positive and 3,634 asymptomatic ANA negative individuals were studied. The mean age and sex distribution among ANA positive and ANA negative individuals were similar across the three genetic ancestries (**Table 1**).

**Table 1.** Demographic characteristics of asymptomatic ANA positive and negative individuals included in this study.

| **Genetic Ancestry\*** | **ANA status** | **Age (yrs) ± SD** | **Female sex, n, (%)** |
| --- | --- | --- | --- |
| **European**<br>(n=3,642) | ANA positive (n=1,236) | 61 ± 16.1 | 865 (70.0) |
| | ANA negative (n=2,406) | 61 ± 15.3 | 1,675 (69.6) |
| **African**<br>(n=859) | ANA positive (n=355) | 56 ± 14.3 | 259 (72.9) |
| | ANA negative (n=504) | 55 ± 13.3 | 351 (69.6) |
| **Admixed American**<br>(n=1,088) | ANA positive (n=364) | 52 ± 14.7 | 281 (77.2) |
| | ANA negative (n=724) | 52 ± 14 | 556 (76.8) |

No statistically significant difference (p-value ≤ 0.05) was observed for age or sex between groups per ANOVA and chi-square testing, respectively. \*Genetic ancestry per the 1000 Genomes Project superpopulation categories as assigned by the random forest classifier. Abbreviations: ANA, antinuclear antibodies, SD, standard deviation, SLE, systemic lupus erythematosus.

After quality controls and imputation, an independent GWAS was performed for each genetic ancestry. As the age and sex distribution was similar across the ANA positive and ANA negative groups, no adjustment for sex and age was needed in the logistic regression models. No genome-wide significant loci were identified in any of the individual ancestries (**Supplementary Figures 2 and 3**). In the multi-ancestry meta-analysis, 6,245,285 SNPs were included. Although no genome-wide significant associations (p-value < 5×10^−8^) were detected, we identified suggestive associations (p-value < 1×10^−5^) across 8 different loci (**Figure 1, Supplementary Table 3**). The most significant association was identified in the SNP rs17211748, a variant upstream of *HLA-DQB1* (p-value = 1.4×10^−6^, OR = 0.82, 95% CI 0.76-0.89). The sensitivity analysis did not show any genome-wide significant association when the cases were restricted to individuals with ANA titer greater than 1:160 in the multi-ancestry meta-analysis (European, n = 138; Admixed American, n = 104; African, n = 60) (**Supplementary Figure 4**). We tested the genetic association within *HLA-DQB1*, i.e. SNP rs17211748, in SLE patients (European, n = 672; African, n = 527; Admixed American, n = 345) compared to asymptomatic ANA negative individuals. We detected a more significant genetic association with rs17211748 in SLE (p-value = 3.7×10^−7^, OR = 1.26, 95% CI 1.15-1.38) compared to asymptomatic ANA positive individuals (p-value = 1.4×10^−6^, OR = 1.22, 95% CI 1.12-1.32) (**Supplementary Figure 5**).

**Figure 1.**
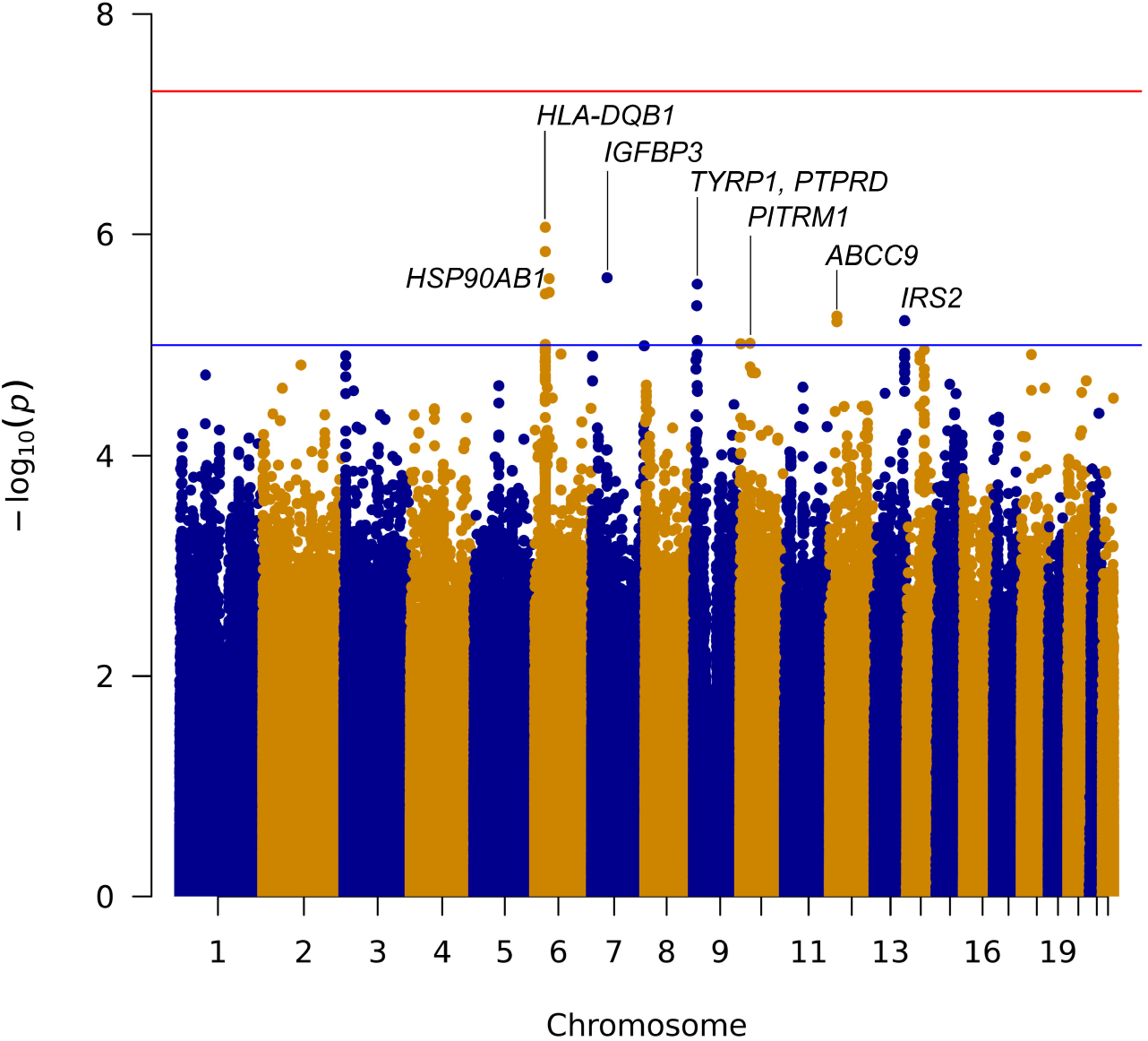
A Manhattan plot depicting the results from the meta-analysis across ancestries comparing asymptomatic ANA positive individuals to asymptomatic ANA negative individuals. The Y and X axes refer to the –log_10_ p-values and chromosome positions, respectively. The red horizontal line represents the genome-wide association threshold (p-value < 5×10^−8^) and the blue line represents the suggestive association threshold (p-value < 1×10^−5^).

To further assess the genetic contribution to ANA production, we calculated a genome-wide SNP-based heritability. Heritability of asymptomatic ANA positivity on the liability scale was estimated to be 24.9% (SD 12.4%).

Finally, we estimated a total genetic risk for SLE in asymptomatic ANA positive individuals compared to asymptomatic ANA negative individuals and to SLE patients. There was a statistically significant difference in the mean SLE GRS between SLE patients and both the asymptomatic ANA positive and ANA negative groups across all three ancestries (**Figure 2, Supplementary Table 4**). No significant difference in SLE GRS between asymptomatic ANA positive and ANA negative individuals was observed in any ancestry (**Figure 2**).

**Figure 2.**
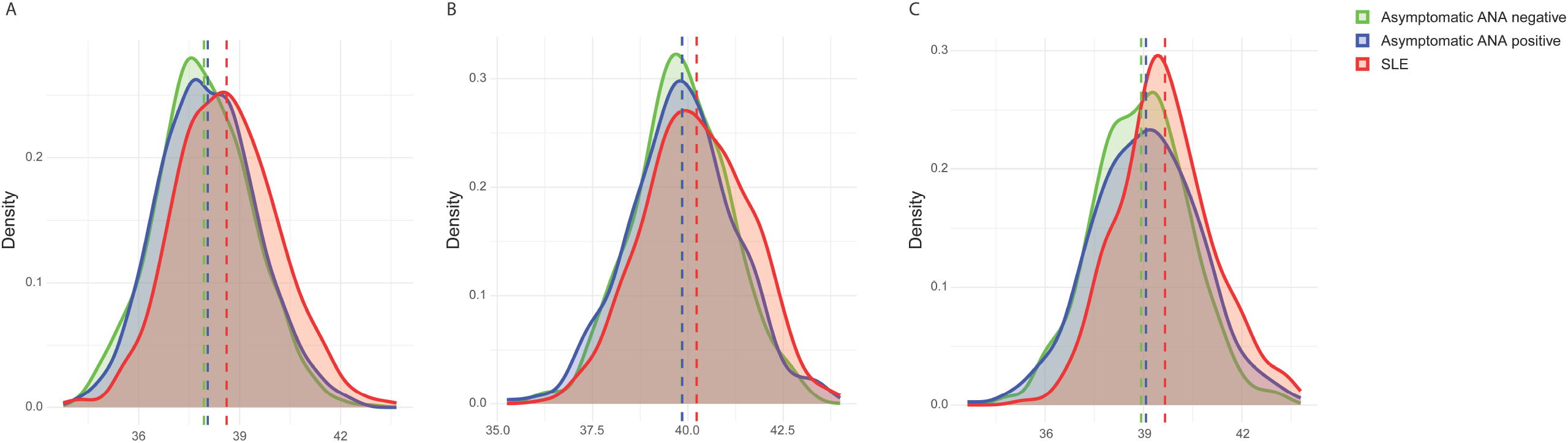
Comparison of cumulative genetic risk scores for SLE in asymptomatic ANA positive individuals, asymptomatic ANA negative individuals, and SLE patients for European ancestry (A) African ancestry (B), and Admixed American ancestry (C). ANA, antinuclear antibody; SLE, systemic lupus erythematosus.

## Discussion

We report a GWAS for asymptomatic ANA positivity across three different genetic ancestries and show that the asymptomatic ANA positive individuals are not enriched for autoimmune genetic risk loci compared to ANA negative individuals, a consistent observation across different ancestries. We estimate the genome-wide SNP-based heritability of asymptomatic ANA positivity to be ∼ 25% suggesting that genetic effects explain only a small fraction of the phenotypic variability due to ANA production in the population. We also show that ANA positive individuals do not exhibit increased SLE genetic risk compared to ANA negative individuals. These results overall suggest that the ANA positivity in the population is primarily influenced by environmental factors.

We did not detect any genetic susceptibility loci in asymptomatic ANA positive individuals with GWAS level of significance. A suggestive genetic association was detected within the HLA class II region, consistent with a previous report in Japanese individuals (5). However, it is conceivable that this genetic effect in our study reflects a small proportion of asymptomatic ANA positive individuals who either have undiagnosed autoimmune disease or are destined to develop autoimmunity in the future. Indeed, the strength of the genetic association signal we detected in *HLA-DQB1*, as reflected by odds ratio, increased when the analysis was performed in SLE patients compared to ANA negative healthy controls.

The role of asymptomatic ANA production as an early event in the pathway to autoimmunity has been well documented (6). Our results show that asymptomatic ANA positivity is primarily driven by non-genetic factors, suggesting that ANA production as an early stage of breakdown of immune tolerance is likely driven by environmental triggers. In a small subset of individuals with sufficient genetic risk for autoimmunity, clinical disease develops. From a disease pathogenesis perspective, our data suggest that while ANA production is essential for the development of SLE, it is independent of the genetic risk for the disease. This raises the potential of developing genetic-environmental models to predict SLE risk in asymptomatic ANA positive individuals. Research on environmental risk factors and gene-environment interactions is needed to build accurate risk prediction models.

This study has several strengths. We report the first multi-ancestral GWAS for asymptomatic ANA production. Ancestry-specific genetic analysis showed consistent observations across genetic ancestries, which increases the generalizability of our results. In addition, we estimated the heritability for asymptomatic ANA positivity for the first time, which we showed to be ∼25%, suggesting a greater role for environmental factors in ANA production.

This study has certain limitations. ANA positivity is a very heterogenous phenotype which includes various autoantibodies directed against nuclear antigens (20). It is possible that the genetic architecture is different across different ANA patterns, titers, or specific autoantibodies. Although we did perform a sensitivity analysis for titers greater than 1:160 and found similar results, our sample sizes were small for this subgroup analysis, limited by the availability of data. As this is a cohort established through EHR, incomplete data or misclassification bias is possible. However, we tried to mitigate this by combining evidence from both clinical codes and medication prescriptions for our case definitions, and setting a very broad definition of autoimmunity. The SNP-based heritability estimate for ANA is based on additive genetic variance only which does not capture all potential sources of genetic variation such as gene-gene and gene-environment interactions.

In conclusion, we performed a multi-ancestral GWAS in a large cohort of asymptomatic ANA positive individuals. We provide evidence to suggest that ANA production is primarily influenced by non-genetic factors and is independent of the genetic risk for SLE. Further research is needed to characterize the environmental determinants of ANA production and the role of gene-environment interactions in the development of autoimmunity in ANA positive individuals.

## Data Availability

All data produced in the present work are contained in the manuscript

