## Supplementary Data and Figures for "Genetic analysis of asymptomatic antinuclear antibody production"

**Supplementary List 1.** Laboratory codes used to define ANA positivity and negativity in All of Us Research cohort.

ANA negative

1. Nuclear Ab [Presence] in Serum by Hep2 substrate (Negative)
2. Nuclear Ab [Presence] in Serum by Immunofluorescence (Not detected/negative, Negative, Not detected)
3. Nuclear Ab [Titer] in Serum by Hep2 substrate (Negative)
4. Nuclear Ab [Titer] in Serum by Immunofluorescence (<= 79, Negative, Not detected, Not detected/negative, Normal)
5. Nuclear Ab pattern [Interpretation] in Serum (Negative, Normal)
6. Nuclear Ab pattern [Interpretation] in Serum by Immunofluorescence (Negative, Not detected)
7. Nuclear Ab pattern [Interpretation] in Serum by Immunofluorescence Narrative (Normal)

ANA positive

1. Nuclear Ab [Presence] in Serum by Hep2 substrate (Positive)
2. Nuclear Ab [Presence] in Serum by Immunofluorescence (Positive)
3. Nuclear Ab [Titer] in Serum by Hep2 substrate (Positive)
4. Nuclear Ab [Titer] in Serum by Immunofluorescence (>= 80, Centromere, Speckled, Nucleolar, Detected, abnormally high, Positive, Abnormal, Present, High)
5. Nuclear IgG Ab [Presence] in Serum by Immunofluorescence (Detected)
6. Nuclear IgG Ab [Titer] in Serum by Immunofluorescence (>=80)
7. Nuclear Ab pattern [Interpretation] in Serum (Speckled, Centromere, Homogenous, Nucleolar, Diffuse)
8. Nuclear Ab pattern [Interpretation] in Serum Narrative (Homogenous, Nucleolar, Speckled)
9. Nuclear Ab pattern [Interpretation] in Serum by Immunofluorescence (Nucleolar, Homogeneous antinuclear antibody pattern, Nucleolar pattern, Centromere, Homogenous, Diffuse, Speckled)
10. Nuclear Ab pattern [Interpretation] in Serum by Immunofluorescence Narrative (Speckled, Homogeneous antinuclear antibody pattern, Centromere, Nucleolar pattern, Nucleolar)

**Supplementary List 2.** List of autoimmune conditions and immunosuppressive medications excluded per SNOMED (Systematized Medical Nomenclature for Medicine) definitions, cohort entry survey and prescriptions in the electronic health records (EHR).

1. Systemic lupus erythematosus
2. Cutaneous lupus erythematosus
3. Other lupus forms
4. Rheumatoid arthritis
5. Inflammatory bowel disease
6. Multiple sclerosis
7. Autoimmune hepatitis
8. Autoimmune thyroid disease
9. Atrophic gastritis
10. Addison disease
11. Immune mediated myositis
12. Scleroderma and related forms
13. Sarcoidosis
14. Autoimmune hemolytic anemia
15. Idiopathic thrombocytopenic purpura
16. Type I DM
17. Sjogren syndrome
18. Vasculitis syndromes
19. Self-reported lupus or rheumatoid arthritis in the survey
20. Methotrexate
21. Leflunomide
22. Mycophenolate
23. Azathioprine
24. TNF-alpha inhibitors
25. Abatacept
26. Apremilast
27. Belimumab
28. Eculizumab
29. Leflunomide
30. Anti-thymocyte globulin
31. Tofacitinib
32. Upadacitinib
33. Anakinra
34. Canakinumab
35. Guselkumab
36. Ixekizumab
37. Sarilumab
38. Secukinumab
39. Tocilizumab
40. Ustekinumab
41. Hydroxychloroquine

**Supplementary List 3**. List of SNOMED (Systematized Medical Nomenclature for Medicine) clinical codes and medication prescriptions used for systemic lupus erythematosus (SLE) case definition.

1. Systemic lupus erythematosus
2. Glomerular disease due to systemic lupus erythematosus
3. SLE glomerulonephritis syndrome
4. Systemic lupus erythematosus encephalitis
5. Secondary autoimmune hemolytic anemia co-occurrent and due to systemic lupus erythematosus
6. Renal tubulo-interstitial disorder in systemic lupus erythematosus
7. Endocarditis due to systemic lupus erythematosus
8. Prednisone
9. Cyclophosphamide
10. Methotrexate
11. Belimumab
12. Voclosporin
13. Azathioprine
14. Hydroxychloroquine
15. Mycophenolate

**Supplementary Figure 1.** Flow chart describing the case and control selection in the All of Us cohort. Abbreviations: AMR, Admixed American ancestry; AFR, African ancestry; EHR, electronic health record; EAS, East Asian ancestry; IFA, immunofluorescence; PC, principal component; SAS, South Asian ancestry.

All of Us participants with genotyping data

(n=312,925)

All of Us participants with genotyping data and no autoimmune disease or medications

(n=254,583)

All of Us participants with genotyping data, no autoimmune disease/medications and ANA test

(n=6,928)

58,342 individuals with autoimmune disease or immunosuppressive medications in the EHR

247,655 individuals with no ANA test by IFA in the EHR

…

Asymptomatic ANA positive

(n=2,563)

Asymptomatic ANA negative

(n=4,365)

Asymptomatic ANA positive

(n=2,211)

Asymptomatic ANA negative

(n=4,051)

352 cases with subsequent negative

314 cases with subsequent positive

EAS, SAS and PC outliers, missing sex, cryptic relatedness removed

EAS, SAS and PC outliers, missing sex, cryptic relatedness are removed

Asymptomatic ANA positive

(EUR 1,236, AFR 355,

AMR 364)

Asymptomatic ANA negative

(EUR 2,406, AFR 504,

AMR 724)

.

**Supplementary Figure 2.** Manhattan plots for the GWAS results across three genetic ancestries: A) European, B) African, and C) Admixed American. Y and X axes refer to the –log_10_ p-values and chromosome positions, respectively. The red horizontal line represents the genome-wide association threshold (p-value < 5×10^-8^) and the blue line represents the suggestive threshold (p-value < 1×10^-5^).

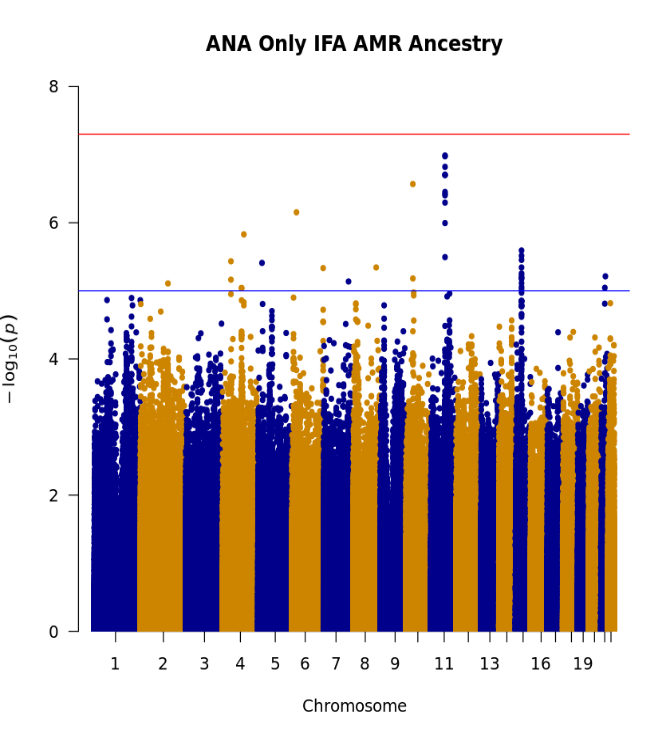

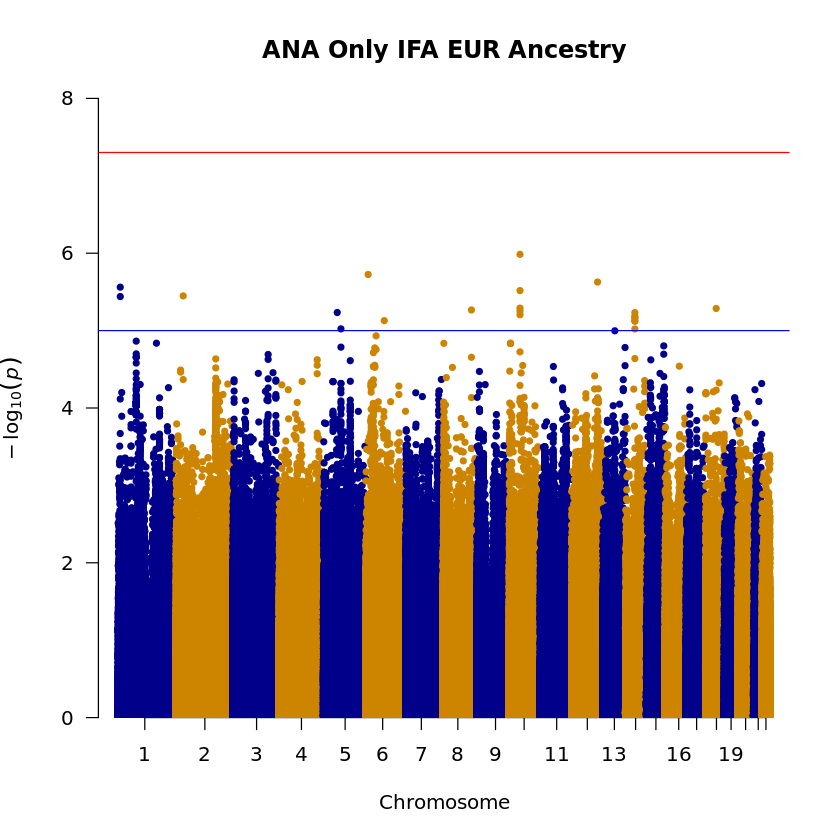

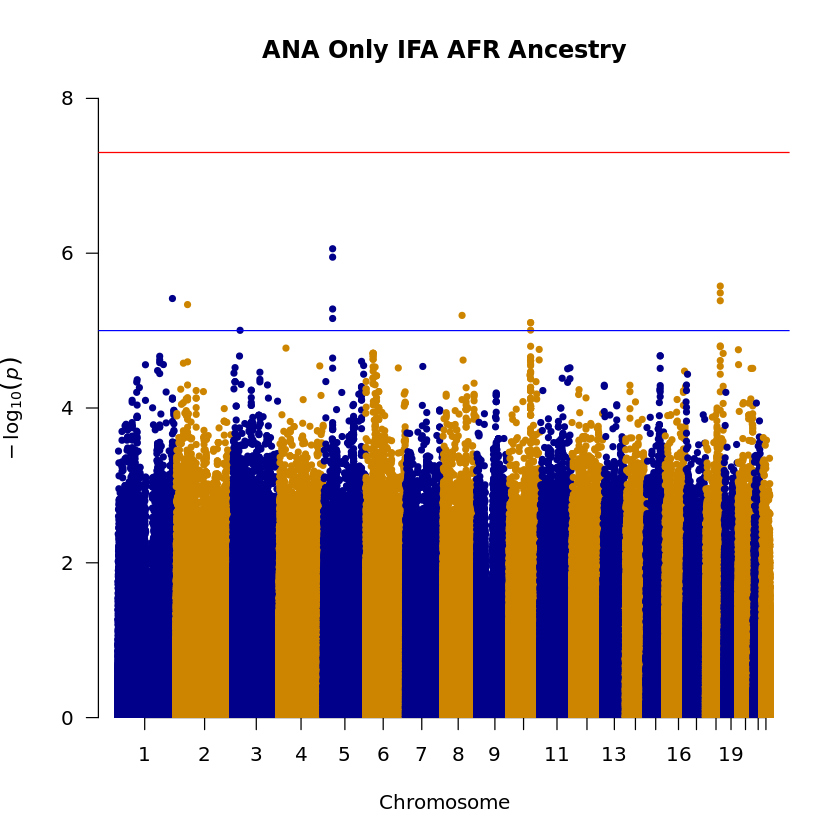

A)

B)

C)

**Supplementary Figure 3.** Quantile-quantile (QQ) plots and genomic inflation factors (λ) of GWAS on individual genetic ancestries: EUR, European, AFR, African, and AMR, Admixed American. The number of single-nucleotide polymorphism (SNPs) included on each study are also indicated.

AMR Ancestry

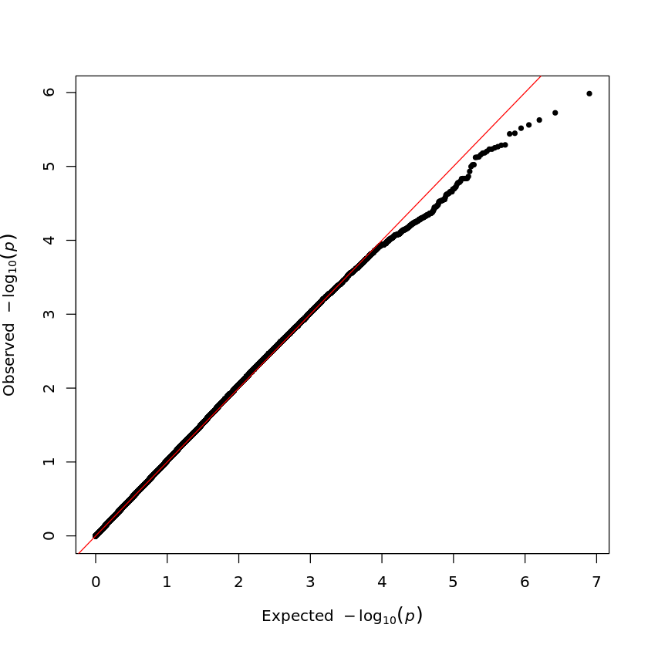

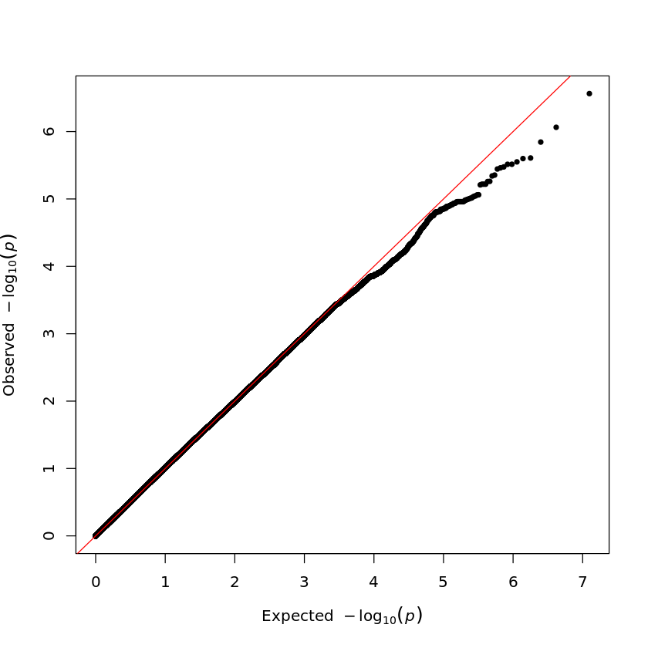

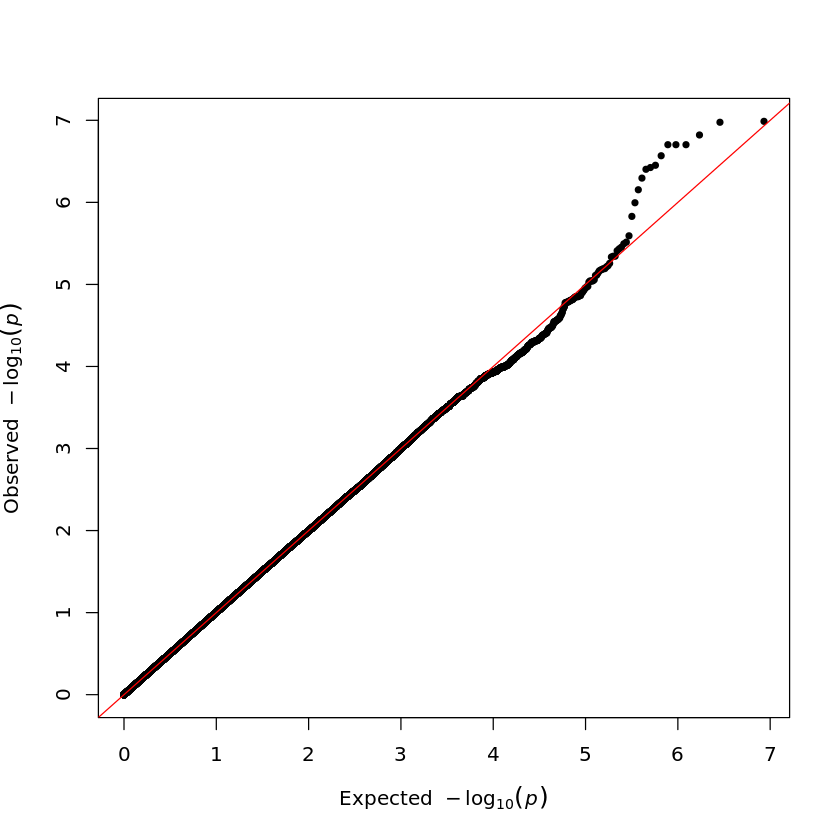

SNPs = 3,960,592

λ = 1.03

SNPs = 5,079,086

λ = 1.02

SNPs = 4,287,233

λ = 1.00

EUR Ancestry

AFR Ancestry

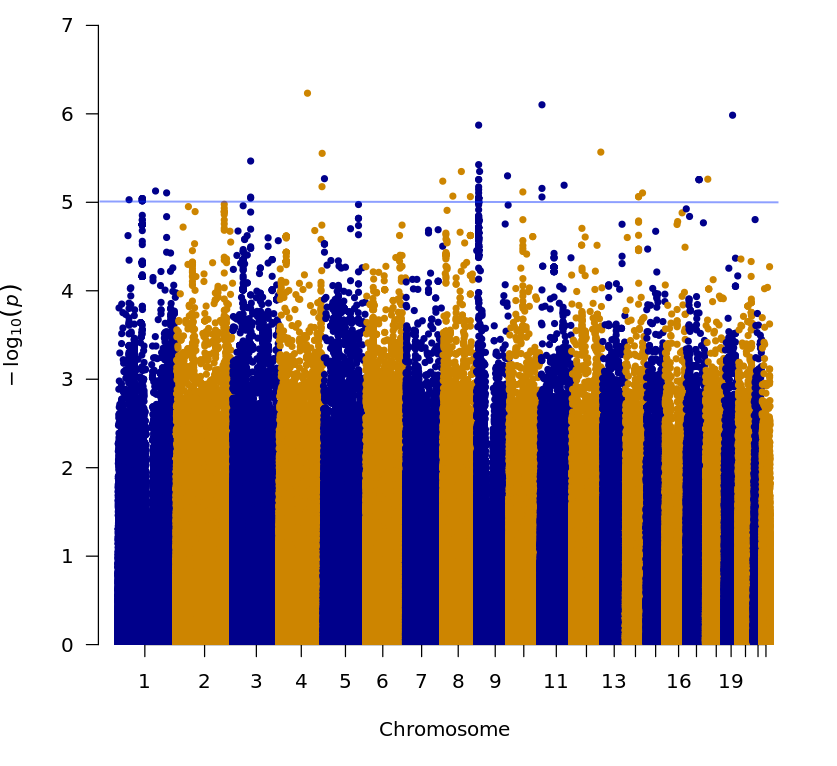
**Supplementary Figure 4.** Manhattan plot depicting the results of the meta-analysis including asymptomatic ANA positive cases restricted to titers greater than 1:160 and asymptomatic ANA negative individuals. Y and X axes refer to the –log 10 p-values and chromosome positions, respectively. The blue horizontal line represents the suggestive threshold (p-value < 1×10^-5^).

**Supplementary Figure 5.** Comparison of odds ratios (OR) and p-values for rs17211148 between SLE, asymptomatic ANA positive, and high titer (≥ 1:160) ANA subgroup. All OR are reported in comparison to asymptomatic ANA negative group. ANA, anti-nuclear antibodies; SLE, systemic lupus erythematosus.

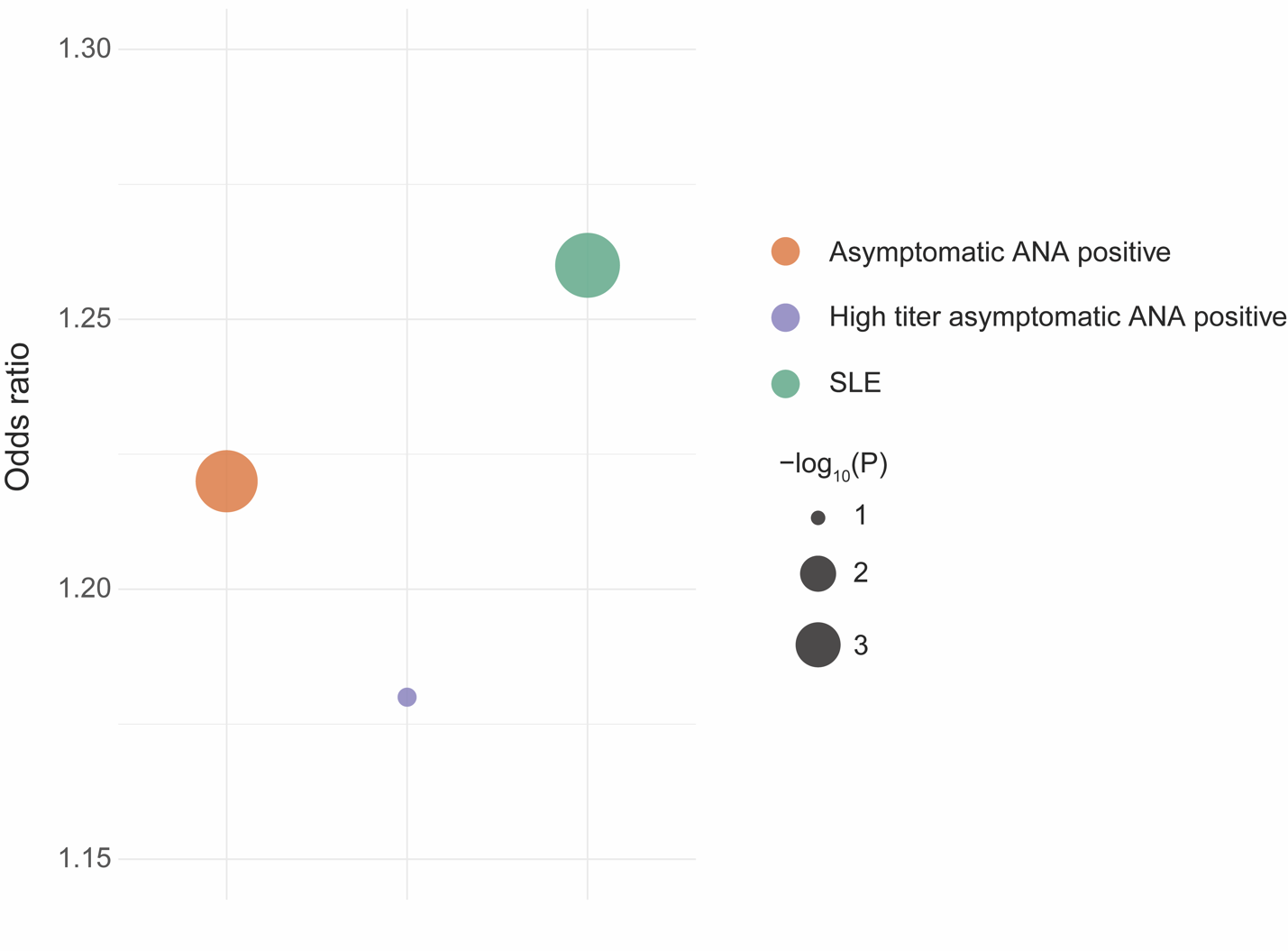
